# Pulsed-Field Ablation Versus Radiofrequency Ablation Combined Ethanol infusion via the vein of Marshall for Mitral Isthmus Line ablation in Persistent Atrial Fibrillation

**DOI:** 10.1101/2025.11.20.25340707

**Authors:** Changyi Li, Lu Zhou, Mengmeng Li, Chenxi Jiang, Xin Zhao, Wei Wang, Ribo Tang, Deyong Long

## Abstract

**Background:** Durable mitral isthmus (MI) bidirectional block is an important substrate endpoint for persistent atrial fibrillation (PeAF) ablation. Radiofrequency (RF) ablation with adjunctive ethanol infusion via the vein of Marshall (EIVOM) improves epicardial lesion formation and MI block durability; pulsed-field ablation (PFA) is a non-thermal alternative that produces rapid myocardial ablation but its acute lesion durability and gap distribution at the MI are not well defined. We compared acute procedural efficacy, lesion durability after a standardized waiting period, ablation time, gap distribution, and safety between PFA and EIVOM+RF for MI line ablation in PeAF.

**Methods:** In this prospective, single-center observational cohort, consecutive patients with symptomatic PeAF undergoing first-time catheter ablation with a planned MI line were enrolled. Analyses included 164 patients (PFA, n=56; EIVOM+RF, n=108). Procedures used standardized mapping and pacing criteria to define bidirectional MI block. Study endpoints include acute bidirectional MI block at the end of ablation, immediate block after MI initial ablation, MI reconnection rate after 20 minutes observation period and total MI ablation time. Residual gap locations and peri-procedural complications were also recorded. Between-group comparisons used appropriate univariate tests.

**Results:** Immediate bidirectional MI block was achieved in 100% of PFA cases versus 97.2% of EIVOM+RF cases (P=0.552). First-pass MI block was significantly higher with PFA (98.2% vs 73.2%; P<0.001). After a 20-minute waiting period, sustained bidirectional block remained in 73.2% of PFA patients versus 89.8% of EIVOM+RF patients (P=0.012). Acute bidirectional MI block at procedure end was 96.4% in PFA group versus 90.7% in EIVOM+RF group (P=0.224). Mean total MI ablation time was markedly shorter with PFA (median 6.1 minutes [IQR 3–8] vs 24 minutes [IQR 16–33]; P<0.001). Residual conduction gaps clustered at the mid-isthmus adjacent to the great cardiac vein in the PFA group (8/13 gaps, 61.5%), whereas gaps in the EIVOM+RF group clustered near the pulmonary vein/ridge region (6/11 gaps, 63.6%). Major complication rates were low and similar between groups (one small self-resolving pericardial effusion in EIVOM+RF; one transient coronary spasm in PFA).

**Conclusions:** In this prospective cohort of PeAF patients, focal PFA achieved very high acute and first-pass MI block rates and substantially reduced MI ablation time compared with EIVOM+RF but demonstrated greater early reconnection after a 20-minute observation period. The two strategies produced different patterns of residual conduction gaps (PFA: mid-isthmus/GCV; EIVOM+RF: PV-ridge), suggesting modality-specific lesion characteristics. These findings support PFA as an efficient approach to MI line creation but indicate that prolonged observation may be required to secure durable MI block with PFA. Further studies assessing longer-term rhythm outcomes and optimized PFA protocols (including potential hybrid approaches) are warranted.

## Background

Recently, some researches have first proved that Mitral isthmus (MI) linear ablation including ethanol infusion into the vein of Marshall (EIVOM) strategies in addition to pulmonary vein isolation (PVI) could significantly reduce atrial arrhythmia recurrence compared with PVI alone. MI ablation is often incorporated into catheter ablation strategies for persistent atrial fibrillation (PeAF). Two main techniques have emerged for achieving durable MI block. The first is conventional radiofrequency (RF) ablation augmented by EIVOM. The vein of Marshall (VoM) lies along the MI and harbors muscular bundles and autonomic fibers; ethanol infusion via the VoM can facilitate epicardial lesion formation to complement endocardial RF. RF ablation alone often fails to produce bidirectional MI block, and reconnections are common, but adjunctive VoM ethanolisation “can facilitate acute MI block” and reduce gap formation[1]. The second approach is pulsed-field ablation (PFA), a non-thermal modality that selectively ablates myocardial tissue while sparing adjacent structures. PFA has demonstrated high safety and efficacy for pulmonary vein isolation in paroxysmal AF and is now being applied to additional left atrial substrates such as the MI[2].

Multiple studies support a significant benefit of adding EIVOM to RF ablation for MI line creation. Previous research found that adjunctive VoM ethanol infusion could raise acute MI block rates as well as shorten required RF time and improved long-term MI line durability. A study showed that durable MI block persisted in 62.9% of the EIVOM group compared to 32.6% with RF alone (p=0.008) in repeat procedures[1]. Prompt-AF and Marshall-Plan trial proved EIVOM+RF strategy could improve long-term freedom from atrial arrhythmias compared with PVI alone[3, 4]. Meta-analyses corroborate these findings: a pooled analysis of >2500 patients found that adding VoM ethanol to AF ablation raised acute MI bidirectional block from 73% to 85% (OR 3.87, p<0.001) and cut atrial tachyarrhythmia recurrence from 42% to 27% (OR 0.52, p<0.001)[5]. In summary, EIVOM adjunct to RF yields markedly higher first-pass MI block rates and better chronic block durability, with resulting improvements in arrhythmia-free survival in PeAF patients. By contrast, PFA offers a novel modality for MI ablation without thermal injury. Early series indicate PFA can achieve high acute MI block, though longer-term data are limited. In the first published experience of PFA for MI in PeAF, Davong et al. reported 100% acute MI block in 45 consecutive patients (all 45 achieved bidirectional block) with infrequent complications. At short follow-up, 20% had recurrent atrial arrhythmia[2]. In a larger prospective cohort, Chen et al. found MI block in 81.3% of patients, with one-year freedom from atrial tachyarrhythmia of 65.6%. Overall PFA procedures (including pulmonary vein and posterior wall ablation) were feasible and safe, with no serious complications like esophageal injury[6]. These studies suggest PFA can deliver effective MI lines in PeAF, achieving block in most cases and acceptable acute outcomes. However, a study showed that only 5.5% patents remained MI block after 3 months MI initial ablation using a PFA catheter[7]. Most of these studies used an over-the-wire PFA multi-spline catheter, which were especially designed to do the pulmonary vein isolation, may not be suitable to ablate the MI line. Previous animal trials suggested that effective lesion formation required of maintains of particularly electrode-tissue contact. Recent research also reported that focal PFA system have a more stabilized tissue contact force, resulting in a non-inferior effect of MI blocking rate and significant less ablation point, which may lead to a higher MI successful ablation rate and a lower risk of complications such as coronary spasm and hemolysis[8]. To our knowledge, our study was the first to focus on verifying the MI ablation effect of a focal contact-force PFA system.

Despite both EIVOM+RF and PFA showing promise for MI ablation, no direct comparisons exist for PeAF patients. The two techniques differ fundamentally: EIVOM uses endocardial RF plus an epicardial ethanol route, whereas PFA uses electric field pulses to produce lesions. These distinctions may influence the distribution of conduction gaps and reconnection sites along the mitral line. Given the strong evidence that EIVOM enhances MI block and may improve outcomes, versus the emerging but still exploratory nature of MI PFA, it is important to compare them head-to-head. A retrospective cohort study focusing on intraprocedural endpoints – such as first-pass bidirectional block rate and locations of residual conduction gaps – will clarify the relative efficacy of these methods. In turn, this analysis will inform how lesion durability might differ between EIVOM+RF and PFA approaches in persistent AF patients. Such comparative data are timely and essential to guide optimal ablation strategies in this population.

## Method

This was a prospective, single-center observational cohort study comparing mitral isthmus ablation using radiofrequency plus ethanol infusion via the vein of Marshall (EIVOM+ RF) versus pulsed field ablation in patients with persistent atrial fibrillation. The study protocol was approved by the institutional review board and all patients provided written informed consent. Consecutive patients with PeAF (defined as continuous AF lasting >7 days) undergoing first-time catheter ablation that included a planned MI ablation line were enrolled. A total of 164 patients between Nov.2024 to Aug. 2025 were included: 108 in the EIVOM+ RF group and 56 in the PFA group. All procedures were performed at a single tertiary electrophysiology center by experienced operators.

### Patient Population

Adult patients (age ≥18) with symptomatic PeAF refractory to at least one antiarrhythmic drug were eligible. Persistent AF was defined according to guidelines as an atrial fibrillation lasting more than 7 days. Key exclusion criteria were prior AF ablation, left atrial thrombus, anatomic contraindications to a mitral isthmus line (e.g. mitral prosthesis), or inability to give consent. Baseline data including age, sex, comorbidities, left atrial size were collected. The two treatment cohorts arose from clinical practice: RF+EIVOM or PFA was performed according to operator discretion and device availability. Baseline characteristics of the groups were compared.

### Ablation Procedure

All ablation procedures were performed under conscious sedation. Vascular access was obtained with femoral venous puncture. Transseptal catheterization into the left atrium was guided by fluoroscopy and intracardiac echocardiography. Unfractionated heparin was administered to maintain an activated clotting time of >300 seconds. A 3D electroanatomic mapping system was used in all cases.

#### EIVOM+ RF group

A multipolar catheter (PentaRay, Biosense Webster) was used for high-density left atrial mapping. First, the vein of Marshall (VoM) was cannulated via a dedicated sheath as described in prior trials. Selective contrast angiography identified the VoM, and 10 mL of 95% ethanol was infused in divided injections into the VoM (ethanol infusion via VoM) to create transmural MI lesions. Following VoM ethanolization, a point-by-point RF ablation line was created from the left inferior pulmonary vein to the mitral annulus using an open-irrigated contact-force RF catheter (ThermoCool STSF, Biosense Webster) with a power controlled mode. A 550 to 600 Ablation index was employed to guide lesion quality in the mitral isthmus ablation procedure.

MI Bidirectional block was checked by differential pacing as described below. The total MI ablation time (sum of RF energy application durations along the MI line) was recorded.

#### PFA group

AForcePlusTM system (Shenzhen Huitai Medical Equipment Co., Ltd, Shenzhen, China), which is a proprietary bipolar PFA system, has been designed for PVI and additional line ablation. The system includes a circular mapping catheter and a contact-force focal PFA catheter embedded with three magnetic sensors allowing model reconstruction, mapping, and ablation in one map with three-dimensional navigation. In brief, a circular mapping catheter from the same system was used for navigation and verification of line formation. The focal PFA catheter was introduced through a deflectable sheath and positioned along the MI path. Point-by-point PFA was delivered along the MI (from the left inferior pulmonary vein ostium to the mitral annulus) with contact-force of 10-15g for each ablation set. The PFA system delivers short trains of high-voltage bi-phasic pulses to induce irreversible electroporation in cardiomyocytes. PFA applications were applied until electrical silence along the ablation path was achieved. As with RF, bidirectional conduction block was assessed after ablation by pacing on either side of the line. All PFA procedures used equivalent energy settings recommended for a focal PFA catheter. No ethanol or thermal energy was used in the PFA group.

In both groups, pulmonary vein isolation was performed as a background procedure if not already present; however, the analysis of MI outcomes was the focus. All procedural parameters (total procedure time, fluoroscopy, etc.) were recorded.

#### MI bidirectional block

MI bidirectional block was confirmed in both groups, according to the following characteristics: 1) proximal-to-distal CS activation pattern when pacing at the left atrial appendage (LAA) or left lateral ridge; and 2) activation from the left atrial lateral wall to the ablation line when pacing at the distal CS or SA interval at LAA is longer when pacing at CSd than pacing at CSp.

### Outcomes

We recorded first-pass MI block: When completing the MI ablation operations of each patient, the MI bidirectional block was immediately tested. If MI block was achieved, the case was classified as first-pass block. If MI block was not achieved at the first-time verification of MI block, additional ablation would be performed to achieve the initial MI block, if after this additional ablation, MI was blocked, this was defined as immediate MI block. Acute bidirectional block was defined by loss of conduction across the line in both directions on differential pacing, using established electrophysiological criteria as described above at the end of the whole procedure. After a 20-minute waiting period without additional ablation (to assess block durability), MI reconnection rate and MI reconnection gap location was recorded. In these patients, residual gap sites were identified by gap mapping with the focal catheter The total MI ablation time was also recorded. There were no clinical follow-up endpoints; only intra-procedural outcomes were analyzed.

### Statistical Analysis

Continuous variables are reported as mean±SD or median (interquartile range [IQR]) depending on normality (assessed by Shapiro–Wilk test), and categorical variables as counts and percentages. Between-group comparisons for baseline and procedural variables were made using χ² or Fisher exact test for categorical data and unpaired t-test or Mann–Whitney U test for continuous data, as appropriate. Mean MI ablation time was compared by weighted t-test (or rank-sum test on weighted data if non-normal). The distribution of gap locations (secondary outcome) was described by group but not formally tested. A two-sided P value <0.05 was considered statistically significant. All analyses were performed using statistical software STATA 17 (Stata Corp. College Station, TX, USA) and followed recommended reporting guidelines.

## Results

### Baseline Characteristics

Table 1 lists baseline demographics and clinical features for the 56 patients in the PFA group and 108 in the EIVOM+ RF group. The two cohorts were well matched: mean age was 63.6 vs 68.5 years, 40.7% vs 39.3% were male. The prevalence of coronary artery disease, hypertension, diabetes, prior stroke, and left atrial diameter, LVEF were also similar between groups (all P>0.05). The CHA2DS2-VA score was similar between the two groups. These findings indicate that baseline demographics and comorbidities were comparable between groups, consistent with prior AF ablation cohorts. Table 1 showed the baseline characteristic of the study.

**Table 1.** Baseline characteristics of the study population. Values are mean (interquartile range [IQR]) or n (%). PFA = pulsed-field ablation; EIVOM+RF = radiofrequency ablation with ethanol infusion via the vein of Marshall. CAD = coronary heart disease; PCI = percutaneous transluminal coronary intervention; CHF = chronic heart failure; UCG = ultrasound cardiogram; LA = left atrial; LVEF = left ventricular ejection fraction.

|  | PFA(n=56) | EIVOM+RF(n=108) | *p* |
| --- | --- | --- | --- |
| Age, y | 68.5(61.5, 76.5) | 63.6(58,71) | 0.997 |
| Female | 22(39.3) | 44(40.7) | 0.869 |
| CHA2DS2-VA Score | 2.1(1, 3) | 1.6(1, 2) | 0.984 |
| Medical history |  |  |  |
| Hypertension | 29(58) | 57(52.8) | 0.603 |
| CAD | 12(24) | 18(16.7) | 0.283 |
| Diabetes | 15(30) | 21(19.5) | 0.157 |
| PCI | 3(6.0) | 7(6.5) | 1.000 |
| Stroke | 6(12.0) | 6(5.6) | 0.197 |
| CHF | 5(9.8) | 10(9.3) | 0.490 |
| UCG parameters |  |  |  |
| LA diameters, mm | 44(41, 47) | 44(41, 47) | 0.435 |
| LVEF, % | 57 | 60 | 0.105 |

### Procedural Characteristics and Outcomes

#### MI blocking rate

Procedural outcomes are summarized below in table 2. Immediate bidirectional MI block was achieved in 100% of PFA patients versus 97.2% of EIVOM+ RF patients (P=0.552). The first-pass MI blocking rate were 73.2% in EIVOM+RF group and 98.2% in PFA group (P<0,001). After a 20-minute waiting period, sustained bidirectional block was present in 73.2% of PFA patients and 89.8% of EIVOM+ RF patients (P=0.012). The acute bidirectional MI block at procedure end was 96.4% in PFA group versus 90.7% in EIVOM+ RF group (P=0.224). The illustration of MI ablation procedure was shown as Figure 1A-D.

**Figure 1A.**
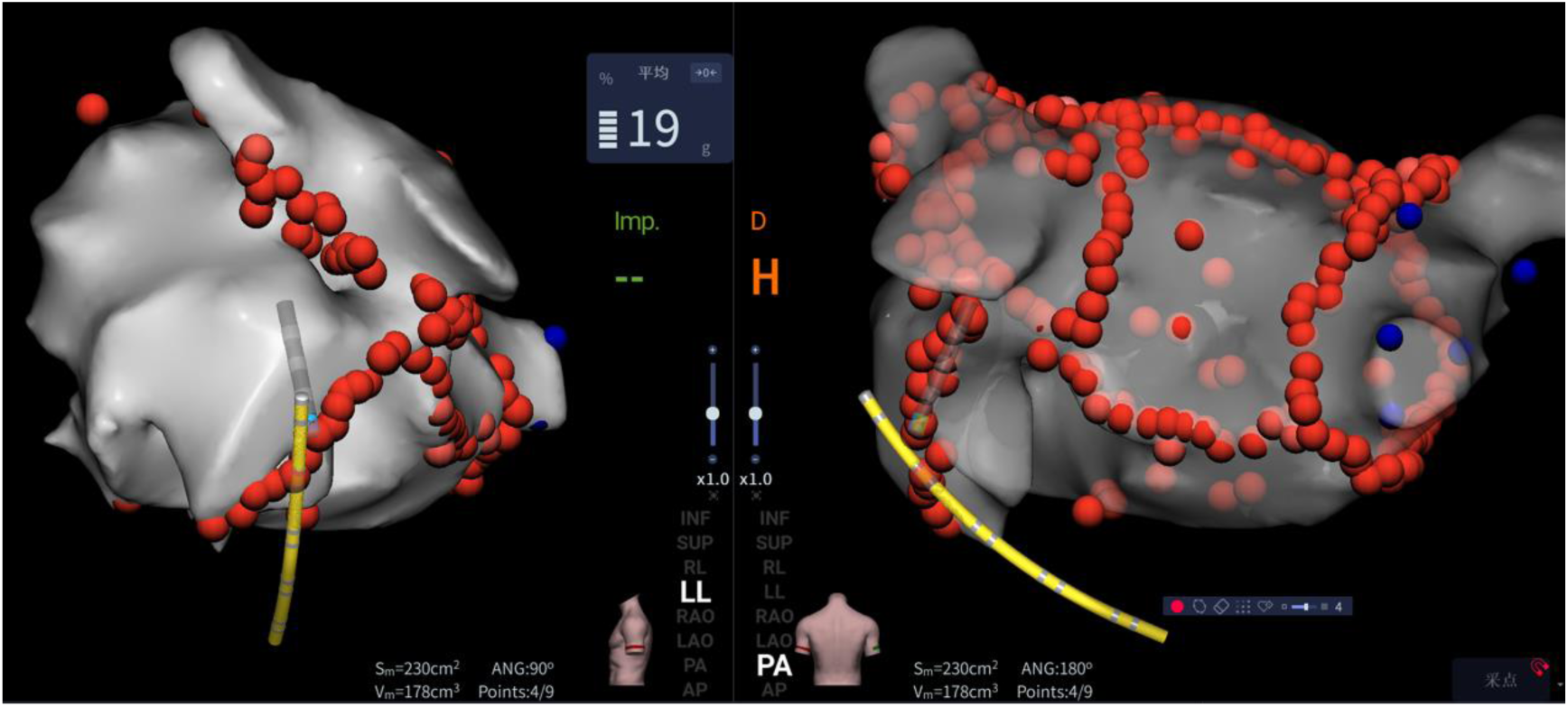
Ablation sets of initial MI ablation

**Figure 1B.**
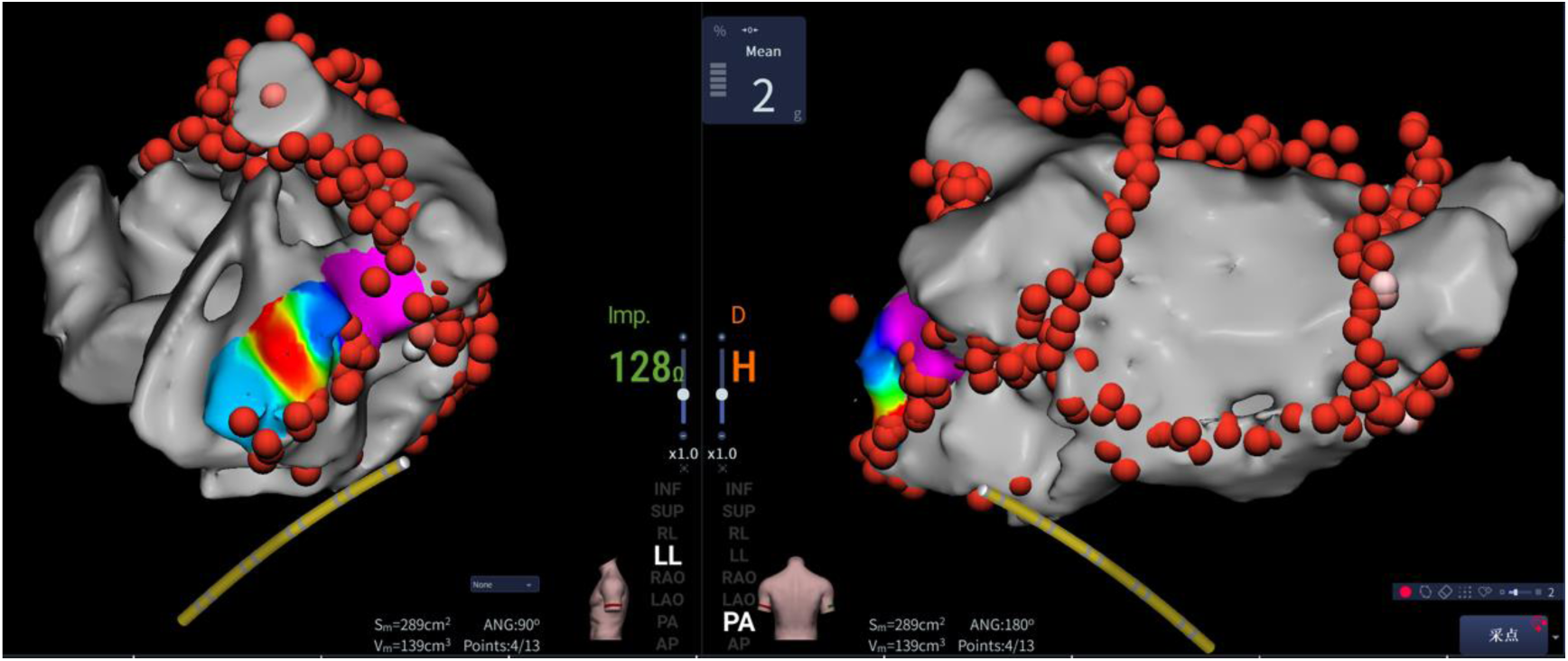
MI reconnection gap location after 20 minutes observation period

**Figure 1C.**
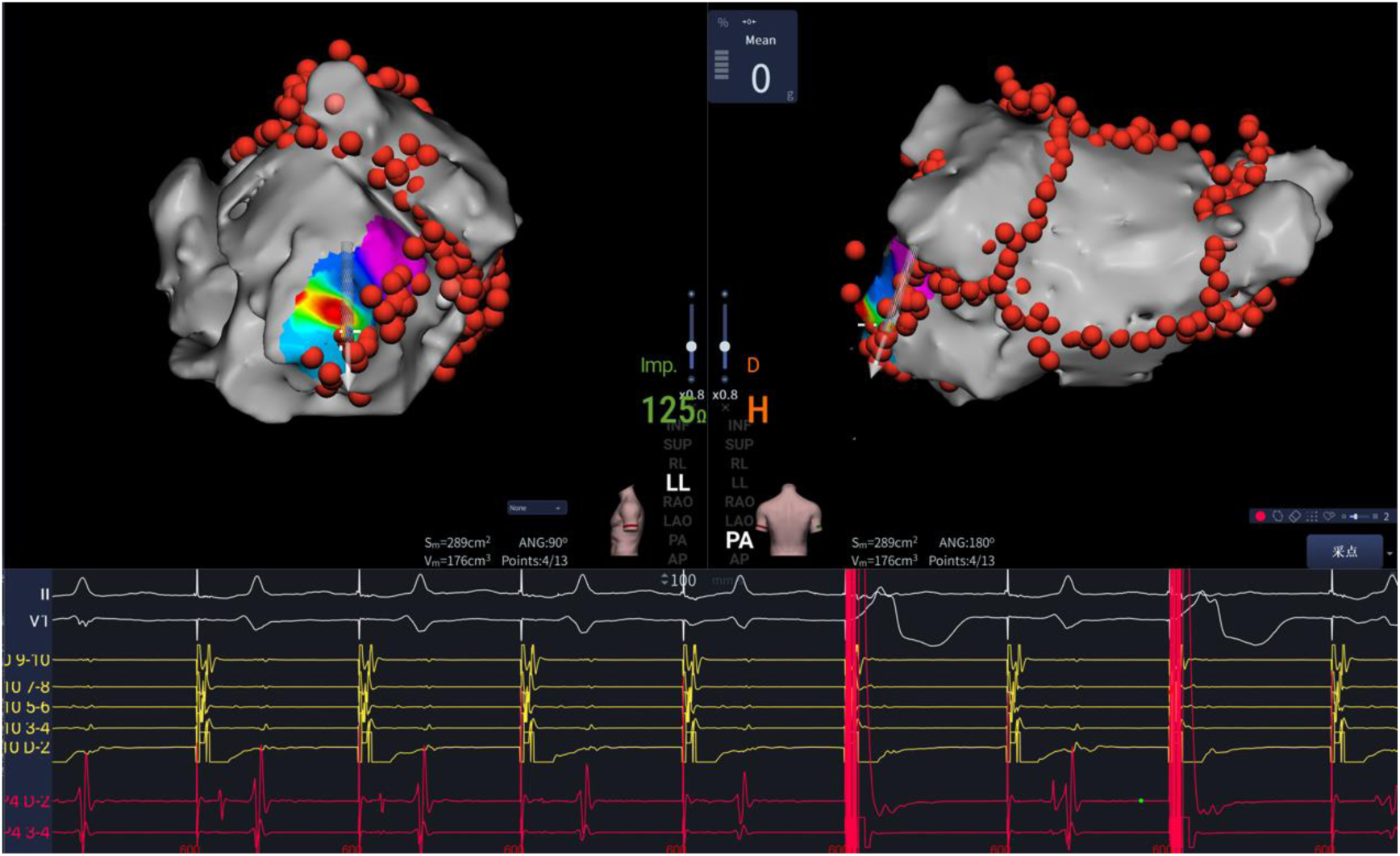
Intracardiac electrogram of MI bidirectional block after additional ablation after MI reconnection.

**Figure 1D.**
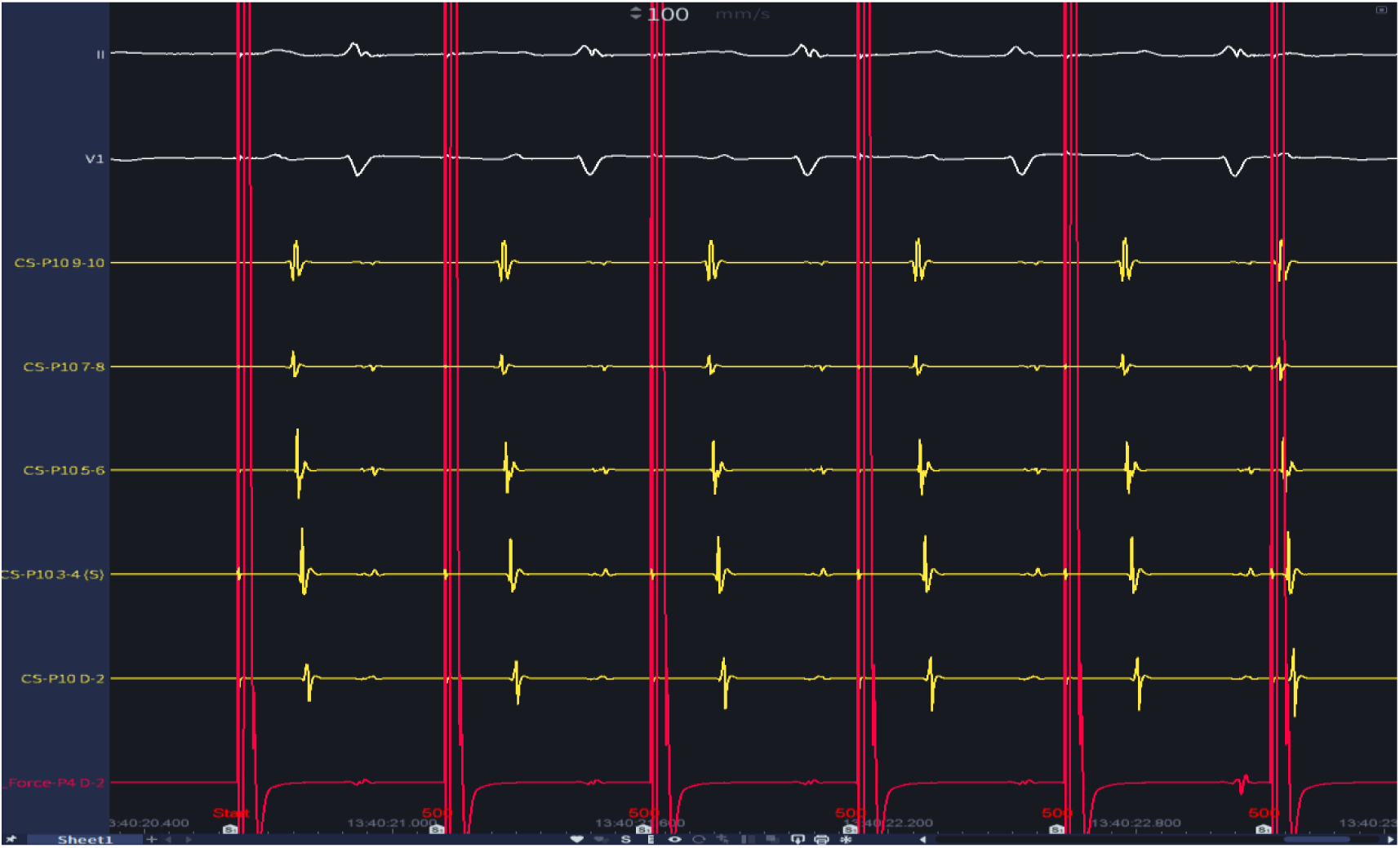
Proximal-to-distal CS activation pattern when pacing at the left atrial appendage after whole MI ablation procedure.

**Table 2.** Ablation characteristics and outcomes. PFA = pulsed-field ablation; EIVOM+ RF = radiofrequency ablation with ethanol infusion via the vein of Marshall. Values are mean (interquartile range [IQR]) or n (%). MI = mitral isthmus

|  | PFA(n=56) | EIVOM+RF(n=108) | *p* |
| --- | --- | --- | --- |
| Total procedure time, min | 6.1(3, 8) | 24(16,33) | <0.001 |
| EIVOM time, min | - | 17.5(6, 24.5) | - |
| MI ablation time, min | 6.1(3, 8) | 6.8(3, 11) | 0.102 |
| Immediate MI bidirectional block, % | 56(100) | 105(97.2) | 0.552 |
| First-pass MI block, % | 55(98.2) | 79(73.2) | <0.001 |
| After 20 min of observation MI recovery, % | 15(26.8) | 11(10.2) | 0.012 |
| Acute MI bidirectional block, % | 54(96.4) | 98(90.7) | 0.224 |

#### MI ablation time

Mean total MI ablation time was statistically lower in the PFA group with 6.1 minutes versus 24 minutes in the combined therapy of EIVOM+ RF group (p<0.001).

### Conduction Gap Distribution

In patients with MI blocking recurrent during the 20 minutes observing period, residual conduction gaps were mapped with a 3-Dimensional mapping system. The vast majority of gaps in PFA groups were located in the midpiece of the MI line (8 of 13 patients, 61.5%) near the great cardiac vein, rather than along the lateral (ventricular) or septal (atrial) side of the isthmus. While the gaps were located mainly near the left pulmonary vein of the isthmus in the EIVOM+ RF group (6 of 11 patients, 63.6%).

### Safety and Complications

Both ablation strategies were safe with low complication rates. In the PFA group, coronary artery spasm occurred in 1 patient (1.7%) without using nitroglycerin. No PFA patient experienced cardiac tamponade, stroke, AV block, or other major injury. In the EIVOM+ RF cohort, 1 patient (0.93%) had a small pericardial effusion (self-resolved). No deaths or cerebrovascular events occurred in either group. Major complication rates did not differ significantly between the two strategies.

Table 2 showed the ablation characteristics and outcomes.

## Discussion

In this perspective cohort study, we found that PFA achieved a higher immediate bidirectional block rate of the MI than EIVOM+ RF. Specifically, acute MI block was attained in 100% of PFA procedures versus 97.2% with EIVOM+ RF, respectively. This finding is consistent with recent reports showing very high acute MI success with PFA. For example, Davong et al. observed 100% acute MI block in 45 patients treated with PFA, and Menè et al. reported 98.3% MI block using a PFA catheter[2, 9]. By comparison, EIVOM+ RF achieve relatively high acute block rates (Marshall-plan trial showed a 93 successful MI block rate, while Prompt-AF study showed an 87.2% MI blocking rate)[3, 4]. Thus, both techniques are effective at first pass MI block, but the PFA may have a modestly higher successful MI blocking rate. Another, we found that PFA could obtain an easier way to block the MI line as the first-pass MI blocking rate were much higher in the PFA group compared to EIVOM+ RF group (PFA VS EIVOM+ RF: 98.2% VS 73.2%).

Despite its high acute efficacy, the durability of the PFA MI lesions was inferior to EIVOM+ RF in the short term. In our series, 20-minute reconnection occurred in 26.8% of PFA lines (drop from 100% to 73.2% block) versus only 10.2% reconnection in the EIVOM+ RF group (97.2%→89.8% block, *p*<0.05). This recovery phenomenon with PFA has not been discovered a clear mechanistic basis. Previous research proved PFA produced almost instantaneous electrogram attenuation due to ionic and osmotic disturbances and edema. Electroporation acutely stuns the myocardium: cell membranes become hyperpolarized and temporarily non - conductive. However, if the delivered electric field is sub - maximal, some cardiomyocytes may recover from reversible electroporation over minutes. As Terricabras et al. note, rapid voltage drop after PFA is driven by edema and cellular stunning, and without sufficiently deep injury some cells “will recover conductivity resulting in reversible electroporation”[10]. Qian et al. similarly explain that reversible electroporation causes a transient loss of conduction (“shock”) that can return after a time[11]. These effects make acute PFA endpoints unreliable unless addressed by longer waiting and touch-up ablation. In practice, extending observation time after MI ablation may be benefited to secure a durable lesion. Our data support this approach: extending extra observing time after ablating the MI line after PFA could convert many of the acute reconnections and this may improve long-term block.

Another notable finding was the difference in residual gap distribution between groups. First, in the EIVOM+ RF cohort, any remaining conduction gaps tended to lie at the lateral or pulmonary-vein (PV) side of the MI ridge. In contrast, the PFA group’s gaps clustered in the mid-isthmus near the great cardiac vein (GCV) region. This pattern likely reflects both anatomic and biophysical factors. After VOM ethanol infusion and endocardial RF, Zuo et al. found that “In the cases that received EIVOM, the recovery gaps were mainly observed in the upper (46.2% [6 of 13]) and middle (30.8% [4 of 13]) regions of MI. In addition, 7.7% (1 of 13) and 15.4% (2 of 13) were in the lower and CS regions, respectively.[12]” In our EIVOM+ RF group, gaps occurred mainly at the PV-ridge (near the left inferior PV), consistent with these classic sites of epicardial breakthrough. By contrast, PFA gaps occurred more centrally along the MI adjacent to the GCV. We think one possible explanation is that the complex tissue composition at the GCV may attenuate PFA energy. PFA energy is highly selective for cardiomyocytes and causes less damage to fat or vascular structures. Thus, when muscle fibers are intermingled with fat and blood vessels (as along the GCV/VOM course), the effective PFA field on the myocardium may be reduced. Any intervening adipose or circulating blood could shunt or absorb the field, blunt lesion depth[13].

To verify this finding, we conducted an animal trial, in brief, 5 healthy Yorkshire swine (57–74 kg) were used in accordance with institutional animal-care guidelines. The same PFA catheter and mapping system as our human study were used in this animal trial. After vascular access, transseptal puncture was performed under fluoroscopic guidance; a steerable sheath was advanced into the left atrium. Left atrial, pulmonary vein and CS/ GCV were reconstruction by the 3-dimentional mapping system. After mapping, the focal PFA catheter was positioned into the left atrial. 3 ablation sets were conducted near the CS/ GCV and 3 sets were conducted on the free-wall of the atrial where far away from the CS/ GCV, with maintaining the same contact-force of each lesion set. All application sites were tagged in the 3D map for later correlation with tissue. The residue steps were completely in line with Hiroshi Nakagawa’s trial[8]. These measurements correspond to the central dark (necrotic) zone seen on TTC. We measured the depth of each lesion and compared the average lesion depth of each group. The results showed that the depth of the lesion near the GCV/VOM were were significantly shorter than those lesions far from the GCV/VOM area (3.74 VS 5.31, p<0.001), the depth of each lesion set were illustrated as Figure 2.

**Figure 2A.**
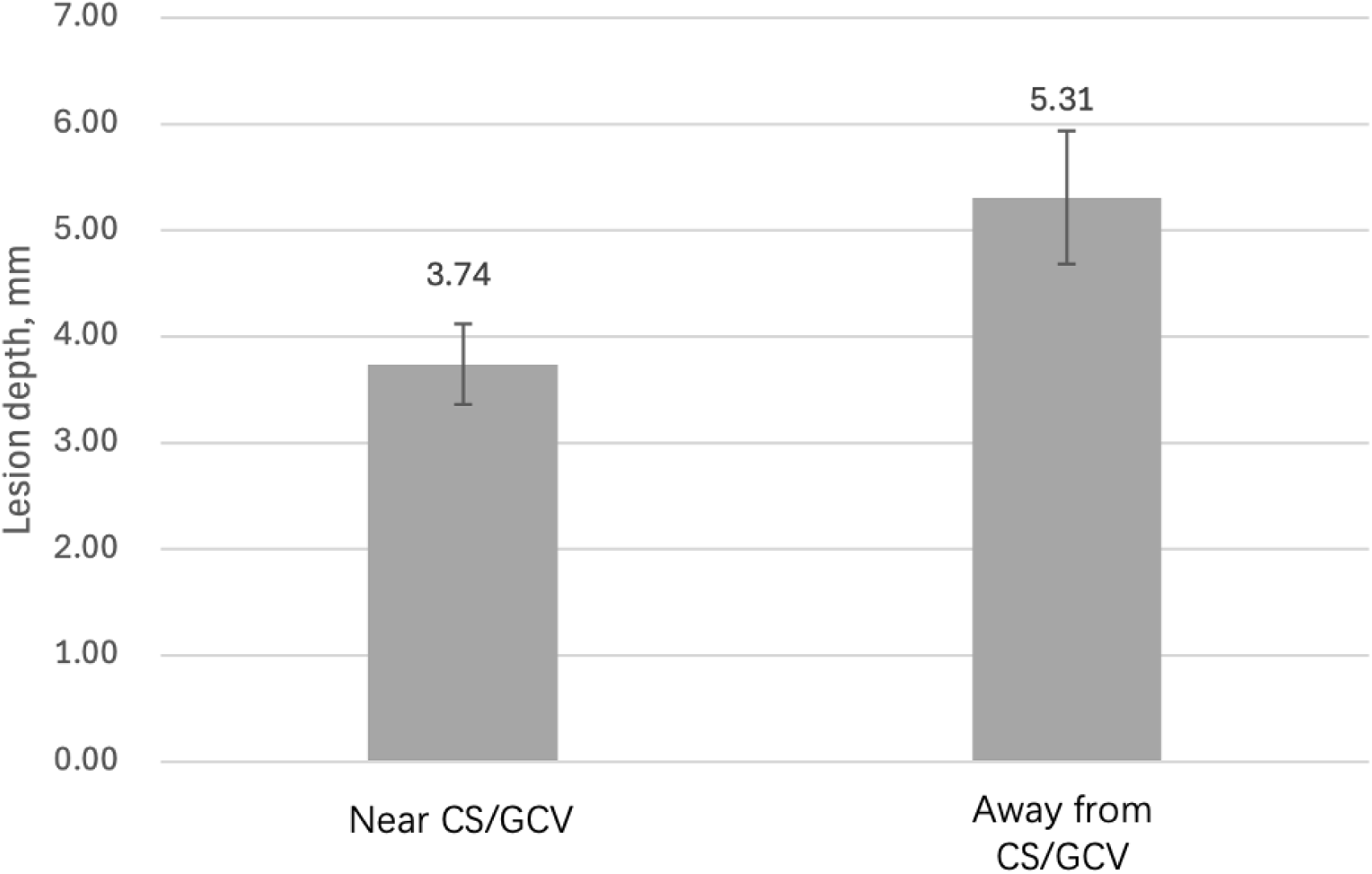
Average lesion depth after triphenyl tetrazolium chloride (TTC) staining in the near the CS/GCV group and away from CS/GCV group.

**Figure 2B.**
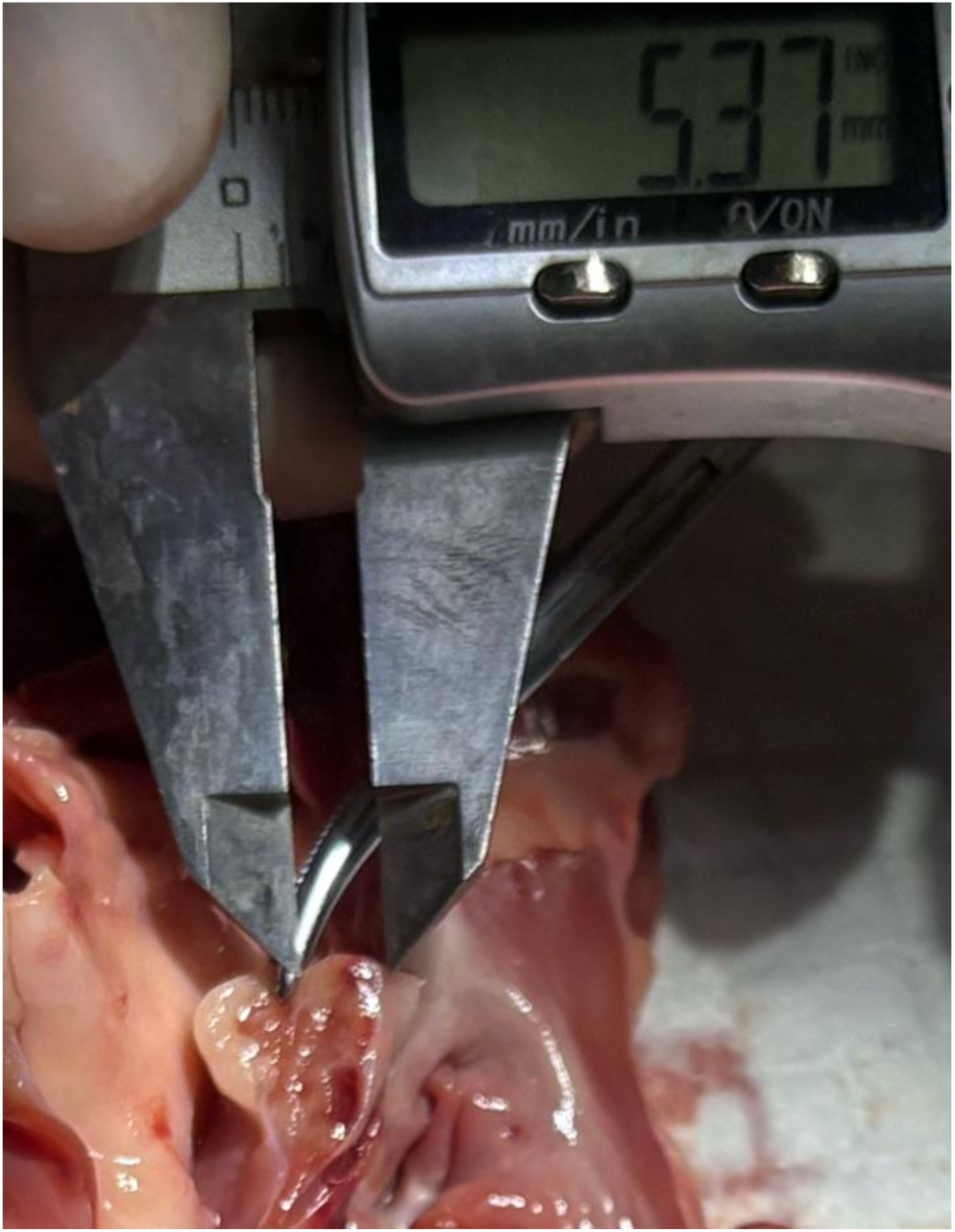
Lesion depth measurement after triphenyl tetrazolium chloride (TTC) staining in the near the CS/GCV group

**Figure 2C.**
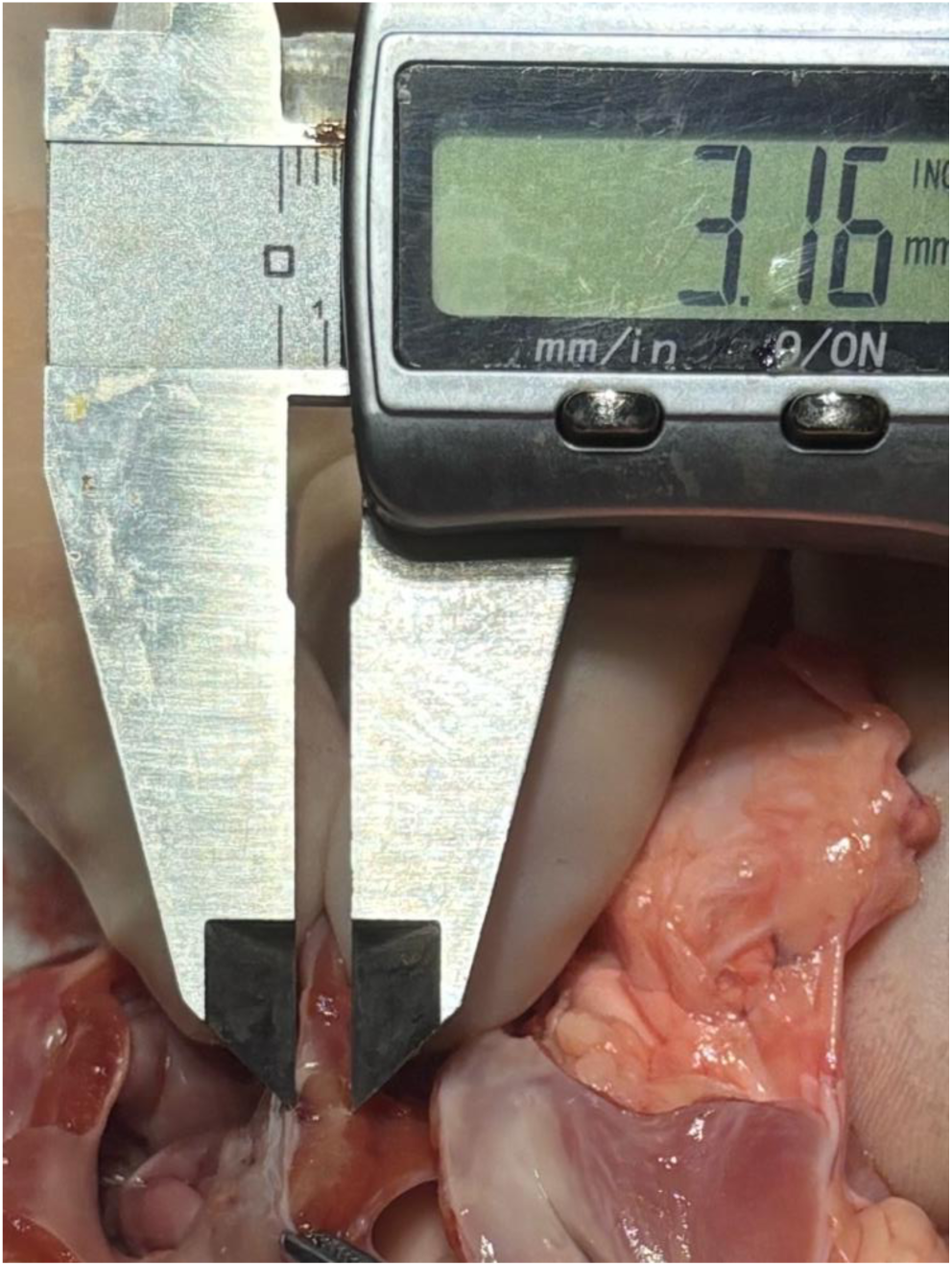
Lesion depth measurement after triphenyl tetrazolium chloride (TTC) staining in the away from the CS/GCV group

In addition, tissue thickness is a known limitation for PFA: thicker myocardium requires stronger fields and may not be fully penetrated by standard PFA protocols. However, previous research proved that the lower 1/3 of MI where near the annular were the thickest, which was diverse to the reconnection location of the MI line in this study[14]. As a result, the tissue thickness may not be the main factor cause MI reconnection of PFA energy. Another perspective believed that the mid-MI region near the GCV may have more epicardial muscle bundles coursing along the vein of Marshall and the adjacent GCV sustain conduction across the MI, so the delivered PFA doses might create incompletely transmural lesions there. We think the reason why the acute MI blocking rate were similar between the PFA group and EIVOM+ RF group may be due to the relatively safety of PFA energy, which would not generate steam pop, may have a lower risk of cardiac tamponade. Consequently, when MI reconnection occurred, the operator may have more confidence to do the additional ablation with PFA energy in a higher position near the ostium of the left atrial appendage opening than the radiofrequency energy.

In summary, the distinct gap patterns between EIVOM+ RF and PFA likely reflect anatomy and the unique physics of PFA energy. Further research may focus on whether the PFA+EIVOM could be more beneficial to the prognosis of atrial fibrillation patients.

Another key finding of our study was that PFA produced dramatically shorter mitral isthmus ablation time than the EIVOM+ RF approach (mean 5.7±5.0 vs. 22±7.9 minutes, p<0.001). This difference is both statistically robust and likely clinically meaningful, the PFA system also required significantly less fluoroscopy and radiation exposure. Especially when using a 3-dimentional mapping system, the fluoroscopy-zero target of ablation MI line could be achieved. These procedural advantages were likely to reducing the immediate and long-term complications caused by radiation exposure both for patient and staff. Notably, our findings echo those in other left atrial ablation settings: for example, the ADVENT trial in paroxysmal AF reported significantly shorter overall procedure times with PFA versus thermal RF, and pooled analyses of PFA versus cryoballoon PVI similarly show much shorter ablation durations with PFA. Together, these data suggest that the markedly reduced ablation and fluoroscopy times seen with PFA are consistent with a general procedural benefit of this technology.

In addition, attention should be paid to safety. Although only 1 patient (1.7%) were experienced transient coronary artery spasm (resolved promptly with nitroglycerin) in our study, the risk of coronary artery spasm during MI by PFA were still an issue of concern. Previous study found that 4.4-41.2% of patients developed acute circumflex spasm without routine prophylaxis strategy[2, 15]. One noticeable reason for the lower risk of coronary artery spasm may be due to the different PFA catheter. In clinical spasm experience, PFA had been performed using a multielectrode pentaspline catheter designed to create a broad electrical field while focal PFA catheter might project a narrower electrical field less likely to cause severe spasm.

These findings have important clinical implications. Firstly, they suggest that contact-force focal PFA alone can be an effective strategy for MI line ablation, but success requires adaptation of technique. Operators should be prepared for high acute block rates but also high acute reconnection. A longer waiting period (e.g. ≥20 minutes) and systematic remapping of the MI line after PFA are warranted. In terms of future research, our results underscore the need for optimization of PFA ablation protocols in the LA. Studies should examine whether higher pulse counts, bipolar vs. unipolar delivery, or adjunctive therapies (e.g. ethanol infusion of the VOM) improve lesion durability. Improved imaging or lesion assessment tools may help verify transmurality in real time. Finally, randomized trials comparing PFA and conventional ablation for persistent AF may include MI line efficacy as an endpoint.

## Limitation

Our study is limited by its focus on acute electrophysiological endpoints. We did not perform >20 min extended observing time after the block of recurrence MI bidirectional MI blocking or conducted the long-term follow-up for rhythm outcomes, so conclusions about sustained sinus rhythm are modest. Davong et al. reported an approximately 20% atrial arrhythmia recurrence rate at 3 months after PFA MI ablation; similar data for our cohorts would be needed. Thus, while we note the potential importance of long-term rhythm control, detailed outcomes will require future study. Other limitations include the relatively small sample size and lack of randomization; nonetheless, the significant differences in reconnection rates are likely robust.

## Conclusion

In conclusion, PFA produces very high immediate MI block rates – higher than conventional EIVOM+ RF – but also a higher rate of conduction recovery. These features of PFA (myocardial selectivity and reversible electroporation) explain the distinct gap distribution we observed and suggest that prolonged observation with extra applications is needed for durable MI lines. Our findings highlight both the promise of PFA and the need to tailor ablation strategy when targeting atrial substrates beyond the pulmonary veins.

## Data Availability

The data that support the findings of this study are available from the corresponding author upon reasonable request.

